# Validation of a simplified HPV genotyping assay designed for cervical screening in low-resource settings

**DOI:** 10.1101/2024.10.21.24315750

**Authors:** Kanan T. Desai, Kayode O. Ajenifuja, Clement A. Adepiti, Federica Inturrisi, Casey Dagnall, Amanda C. Hoffman, Didem Egemen, Julia C. Gage, Nicolas Wentzensen, Silvia de Sanjose, Mark Schiffman

**Affiliations:** National Cancer Institute, Rockville, MD, USA; Norwalk Hospital Internal Medicine Residency Program, Norwalk, CT, USA; Obafemi Awolowo University, Ile-Ife, Nigeria; Cancer Genomics Research Laboratory, Frederick National Laboratory for Cancer Research, Leidos Biomedical Research, Inc., Frederick, MD, USA; ISGlobal, Barcelona, Spain

**Keywords:** Cervical cancer screening, HPV testing, HPV genotyping, point-of-care, LMICs

## Abstract

**Introduction:** Human papillomavirus (HPV) genotype predicts cervical cancer risk, and genotyping could help guide management of HPV positives as part of cervical screening. An isothermal amplification HPV extended genotyping test (ScreenFire HPV RS assay) can assay up to 96 controls/samples in one hour plus preparation time. A novel format with pre-aliquoted reagents and an anti-contamination component (Zebra BioDome™) could simplify the HPV testing process by substantially reducing the assay preparation time and the chances of post-amplification contamination. We validated Zebra BioDome formulation prior to its clinical use.

**Methods:** Residual provider-collected cervical samples (n=450) from a population-based study in rural Nigeria were retested with ScreenFire, once using the standard assay version (liquid reagents combined onsite) and twice with Zebra BioDome. HPV results with adequate DNA (N=427) were analyzed channel-by-channel and using the cervical cancer risk-based hierarchy of HPV type channels (HPV16, else 18/45, else 31/33/35/52/58, else 39/51/56/59/68, else high-risk HPV negative) to evaluate Zebra BioDome repeatability and accuracy against the standard version.

**Results:** Zebra BioDome reduced the number of pipetting steps to run the ScreenFire HPV assay. Following amplification, the BioDome material formed a sealant layer above the reaction components. Zebra BioDome had excellent repeatability and agreement with the standard version, both at the channel-specific analysis [positive percent agreement between 88.4% (HPV39/51/56/59/68) and 100% (HPV16); negative percent agreement between 97.8% (HPV31/33/35/52/58) and 100% (HPV39/51/56/59/68)] and hierarchical analysis (overall agreement 97.2%).

**Conclusions:** The assay version utilizing Zebra BioDome performed similarly to the previously validated standard version of the ScreenFire HPV assay and is now undergoing field evaluation. This solution has the potential to reduce assay preparation time and risk of contamination, providing a simpler, low-cost, near-point-of-care HPV testing and extended genotyping solution for cervical screening in lower-resource settings. Potential application of Zebra BioDome technology to other DNA amplification assays should be considered.

## Introduction

Cervical cancer rates and deaths tend to be highest in regions with limited resources (1). The World Health Organization (WHO) call to eliminate cervical cancer globally promotes human papillomavirus (HPV) vaccination, HPV-based screening during mid-adulthood, and treatment of both precancerous and cancerous conditions (2). Highly sensitive HPV testing of self-collected samples is being promoted worldwide (3), because of the crucial role of HPV in the development of cervical cancer (4), and the pressing need to screen many millions of women. It is impractical to treat every infected woman in high-prevalence areas, because HPV infections are extremely common and often benign. Most HPV infections are controlled by the immune system within 1 to 2 years of detection, and only persistently detectable infections lead to precancer and cancer (4). Because HPV detection on population screening can be very common, and follow-up to distinguish persistence is often impractical, a triage test to identify biomarkers of persistent/progressive infections is necessary for prioritization of care by need. HPV typing is a promising means of triage, because the likelihood of progression depends on the HPV type and varies significantly among the 13 high-risk HPV types (5,6) classified by the International Agency for Research on Cancer (IARC) as Group 1 (known carcinogens) or Group 2A (probable carcinogens) (7). Risk-based extended HPV typing is proposed to stratify HPV-positive women according to their risk of developing cervical cancer and to guide clinical management (6).

An isothermal DNA amplification technology-based ScreenFire HPV risk stratification (RS) assay (Atila Biosystems Inc., Sunnyvale, CA, USA) can provide extended genotyping for 13 high-risk HPV types in the four established risk channels: 16, 18/45, 31/33/35/52/58, and 39/51/56/59/68 (8,9). It was designed to support clinical management according to the risk of cervical cancer associated with the grouped types, hence the emphasis on risk stratification, in which the highest-risk positive channel is reported when multiple concurrent infections are present. It is relatively rapid and low-cost, making it suitable for extended HPV genotyping beyond high-resource settings. Its clinical accuracy has already been shown (8–11) and the assay is currently undergoing field validation as part of the HPV-Automated Visual Evaluation (PAVE) protocol in nine low- and middle-income countries (LMICs) worldwide (12). Two aspects of performing the ScreenFire HPV RS assay emerged during the validation to date and are addressed here: the need to further decrease the time and manual handling needed to run the assay, particularly during assay preparation phase, and the risk of laboratory contamination. Despite being a simple assay, the ScreenFire HPV RS test in its current format still requires that a trained laboratory technician carry out a number of preparatory steps: thawing reagents, preparing the master mix (by combining reaction mix and primer mix), dispensing the master mix into reaction tubes, and then adding the sample (Figure 1A). These steps involve pipetting small volumes of reagents and samples, demanding careful attention to detail and increasing the risk of human error, cross-contamination, and variability in results. Moreover, these steps add to the overall time required for HPV testing, making single-day screening strategies more difficult.

**Figure 1:**
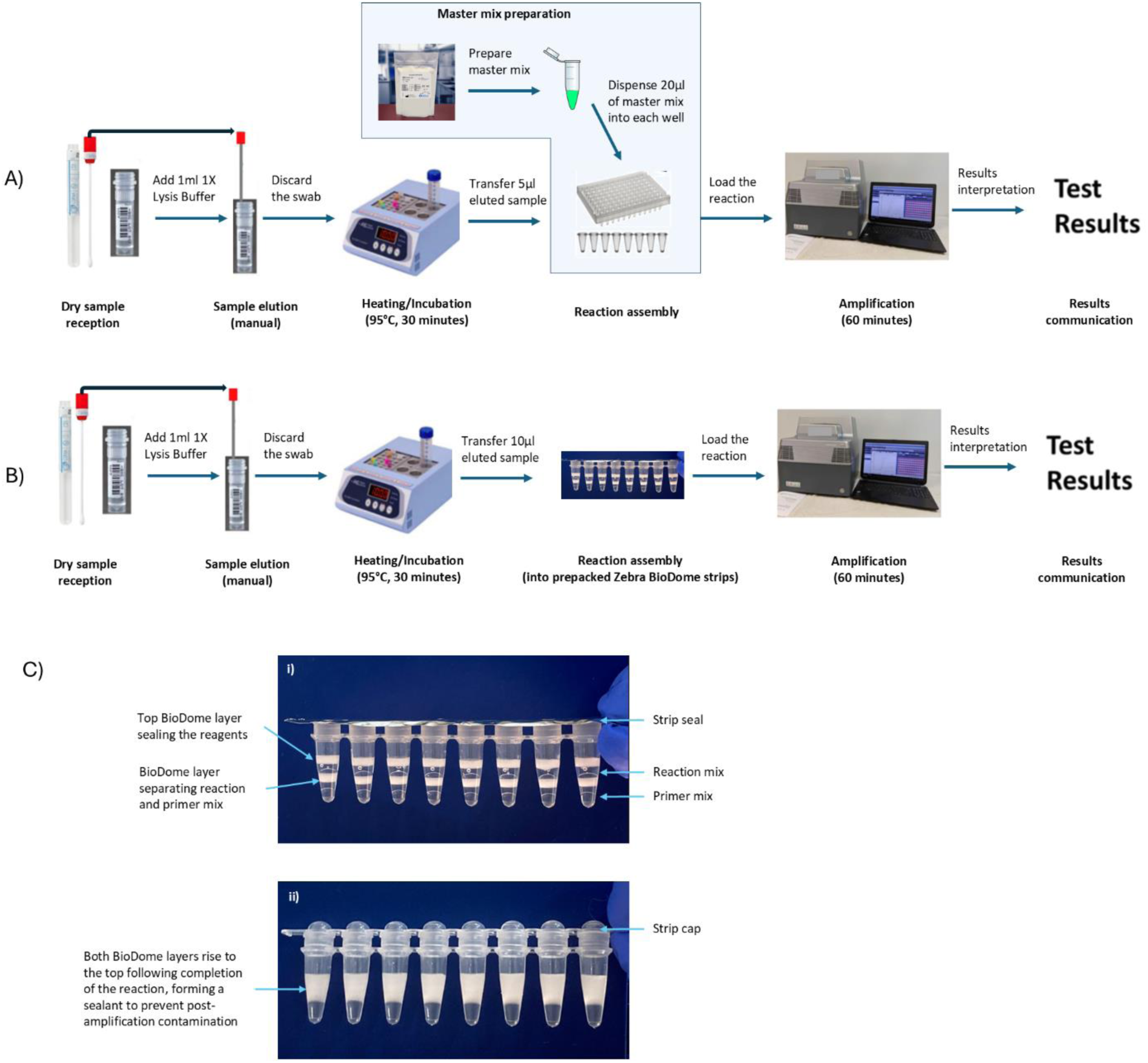
Steps to run the ScreenFire HPV RS assay on the Powergene instrument using dry swab samples following the PAVE study protocol (12). A) Assay in its standard liquid version (the blue box highlights the steps impacted by the introduction of Zebra BioDome). B) Assay in the novel Zebra BioDome version. C) Details of the Zebra BioDome version before (i) and after (ii) amplification.

Moreover, like any other DNA amplification technique (13), ScreenFire HPV RS assay is susceptible to contamination of the laboratory environment by amplified products. The problem arises because the post-amplification concentration of amplicons in a positive sample/control can reach 100-1000 billion copies per microliter. If the tube content is not strictly contained after amplification, the high concentration of amplicons can contaminate laboratory surfaces, equipment, pipettors, and personal protective gear like gloves and laboratory coats. Identifying and cleaning up this contamination is time-consuming and resource-intensive, both in terms of labor and money, and it precludes HPV testing activities until full decontamination has been verified.

To reduce the number of pipetting steps and mitigate the risk of contamination, the assay’s manufacturer Atila BioSystems developed an assay format with pre-aliquoted reagents and an anti-contamination component, known as ‘Zebra BioDome™’. This format removes the need for preparation and dispensing of the master mix, only requiring the addition of the lysate to the Zebra BioDome tubes (Figure 1B) as the two components of the master mix are suspended between gel matrix layers. The gel matrix melts during amplification enabling the mixing of sample and reagents and solidifies again at the top of the reaction volume during cooling, thus creating a physical barrier for the amplified products (Figure 1C).

This article aims to independently validate the Zebra BioDome version of the ScreenFire HPV RS assay by comparing it with the established standard liquid version of the assay and assessing its repeatability.

## Materials and Methods

### Study population and design

The present study drew from 1,339 residual cervical samples stored frozen in PreservCyt solution (Hologic, Marlborough, MA, USA), collected by providers from a population-based screening project in rural Nigeria (Project Itoju) involving women aged 16 to 88 years (mean [standard deviation] = 44.0 [15.8]). The methodology of the Project Itoju has been detailed elsewhere (14). In brief, between 2009 and 2010, approximately 1,420 eligible women—who were not pregnant, were sexually active, had not undergone a hysterectomy, were at least 15 years old, and had been residing in their household for more than 3 months—were invited from homes in the village of Irun, Nigeria. Participants in the research effort provided written informed consent. During the screening, locally trained nurses conducted cervical examinations and collected samples using a broom-type device and an endocervical brush, which were then placed in PreservCyt for liquid-based cytology (LBC) and HPV testing.

One milliliter aliquots of the frozen residual cytology samples were tested in the United States (Albert Einstein Cancer Center) for HPV using an AmpliTaq Gold MY09-MY11 polymerase chain reaction (PCR)-based test that included additional type-specific primers, and primers to amplify a cellular beta-globin fragment as an internal control for amplification as previously described (15). PCR products were genotyped with dot-blot hybridization using type-specific probes for 13 high-risk and >20 other HPV types, using a semi-quantitative measure of signal strength (1-5 from lowest to highest) as a proxy of viral load (16). Of note, the PCR signal strength variable (1–5) is not strictly linear, in that a signal strength of 5 has no upper bound and is, on average, particularly strong. Samples were also tested for HPV16 and HPV18 using a research-use-only (RUO) HPV DNA test, the Luminex assay.

Out of the 1,339 residual samples, valid (i.e., adequate DNA) HPV results at the AmpliTaq Gold MY09-MY11 PCR were obtained for 1,305 samples. A small subset (n = 8) was unblinded and sent to Atila BioSystems for updating the AmpFire assay chemistry to the ScreenFire RS HPV assay with its emphasis on sensitive detection of the most high-risk HPV types (8). Of the remaining 1,297 masked samples with valid results, we selected all 356 samples that tested positive for any HPV types (including low-risk types) by either PCR or HPV16/18 Luminex.

Additionally, we randomly chose 100 of 941 specimens that tested negative for all HPV types (including low-risk) by both PCR and Luminex. These 456 samples were previously used to redesign and validate the ScreenFire HPV RS assay in its current standard format (8). Out of the 456 samples, six were found to have insufficient residual material (i.e., no more volume), leaving 450 usable samples.

These same 450 samples were retested with ScreenFire HPV RS assay using both the standard (9) and Zebra BioDome versions of the assay. Testing using the standard version was conducted at the US National Cancer Institute’s (NCI) Cancer Genomics Research (CGR) laboratory with visiting Atila BioSystems laboratory scientists. Testing using the Zebra BioDome version was conducted at the Atila BioSystems Laboratory by Atila BioSystems scientists. The scientists were blinded to prior testing results and had no role in the analysis or reporting of the experiments.

### ScreenFire HPV testing

As per manufacturer’s protocol, ScreenFire HPV RS assay can be used with dry swab samples, cervical cell suspension, FFPE slice samples, and purified DNA samples. For the work presented here, the protocol for cervical cell suspension was used, plus pre-processing steps to prepare the frozen residual specimens. Initially, 1 mL of cervical cell suspension in PreservCyt was transferred to a 1.5 mL tube and centrifuged at maximum speed for 15 minutes. The supernatant was discarded, and 100 μL of 1X lysis buffer was added to the pellet. The mixture was vortexed thoroughly to resuspend the cells. The resuspended cell pellets were then transferred to 96-well PCR plates, incubated at 95°C for 15 minutes, allowed to cool to room temperature, and stored at −20°C for future use.

The plates were thawed at room temperature prior to testing with ScreenFire HPV RS assay. This was done twice on one Powergene 9600 real-time PCR machine, once using the standard liquid version and once using the Zebra BioDome version, and once on a second Powergene machine using the Zebra BioDome version.

For testing with the standard liquid version, 5 μL of the prepared specimen (lysate) was combined with 20 μL of freshly prepared master mix (containing reaction mix and primer mix) in a 96-well PCR plate using a pipette. For testing with the Zebra BioDome version, 10 μL of the lysate was added to each well using the technique described below. Positive and negative template controls were included in each run. The plates or tube strips were sealed or capped, gently vortexed, and centrifuged to ensure that all liquid settled at the bottom of the wells.

The plates or Zebra BioDome tube strips were then placed into the Powergene 9600 real-time PCR machines, set to the isothermal program mode (60°C for 60 cycles of 1 minute each). Fluorescence signals were detected using CY5 for the HPV16 channel, ROX for the HPV18/45 channel, CY5.5 for the HPV31/33/35/52/58 channel, FAM for the HPV39/51/56/59/68 channel, and HEX for the human beta-globin gene as the internal control channel. A sample was deemed positive for an HPV type (group) if a signal was detected in the corresponding HPV channel within 60 minutes, regardless of the HEX channel signal. If no signal was detected in any of the HPV channels within 60 minutes, a signal in the HEX channel was required for a certain sample to be considered a valid HPV negative. If no signal was detected in any of the channels including the HEX channel, a sample was considered invalid suggesting inadequate DNA in the reaction.

### Zebra BioDome technology

Zebra BioDome is a proprietary format for isothermal amplification assays by Atila BioSystems. It features layers of reaction mix and primer mix separated by the BioDome anti-contamination gel matrix, with an additional BioDome layer on top for stability and containment. This assay format comes as prepacked reaction tube strips covered by an adhesive film to prevent evaporation (Figure 1C-i).

The version of Zebra BioDome described here is penetrable, meaning that at room temperature, the gel matrix is semi-solid – solid enough to prevent loss of the reaction mix, but soft enough to be pierced with a standard pipette tip. Users only need to remove the adhesive film and pipette the lysate into the reaction tube. A standard volumetric pipette with a disposable tip (avoiding very narrow tips to prevent gel matrix blockage) is used to insert the sample/control to the bottom of the Zebra BioDome tube. Once the pipette tip touches the bottom, the pipettor is slightly withdrawn to allow dispensing of the lysate within the primer mix layer. The pipette is then removed and visually inspected to ensure no clogging of residual liquid in the tip. If clogging occurs, the volume transfer of 10 μL lysate is repeated into the same Zebra BioDome tube.

The gel matrix melts during amplification at 60°C enabling the mixing of the reaction components and re-solidifies at the top of the reaction during cooling, thus creating a physical barrier for the amplified products (Figure 1C-ii) and resulting in a sealed reaction chamber.

### Statistical analysis

Valid ScreenFire HPV results were obtained for 427 out of 450 samples for both the standard and Zebra BioDome versions, which were included in the analysis. The remaining 23 were excluded from the analytical sample because of invalid results on either or both assay versions (7 invalids by Zebra BioDome, 7 invalids by standard, 9 invalids by both).

Analyses were done to assess the repeatability of Zebra BioDome when testing on two different Powergene instruments and to compare the HPV results of Zebra BioDome to those of the standard assay version.

Two types of analyses were conducted. First, channel-by-channel in a non-hierarchical fashion, acknowledging that a sample could test positive for multiple HPV channels. Then, hierarchically using the risk-based grouping of HPV genotype channels intrinsic to ScreenFire’s design. In the hierarchical analysis, HPV results were categorized as HPV16 positive, otherwise positive for HPV18/45 (if HPV16 was not detected), otherwise positive for HPV31/33/35/52/58 (if HPV16/18/45 were not detected), otherwise positive for HPV39/51/56/59/68, and otherwise negative for the 13 high-risk types.

Positive and negative agreement statistics were calculated, with 95% confidence intervals (CI) estimated under normal approximation. Agreement statistics for the repeatability analysis were calculated based on either replicate positivity/negativity. Agreement statistics for the comparison of Zebra BioDome against the standard version were calculated using the standard assay version as the reference. Additionally, McNemar’s test for asymmetry was computed for the channel-specific analysis. The analyses were adjusted back to the original sample by weighting HPV negatives (i.e., 100 negatives were weighted by 9.41 for 941 negatives in the original sample) while retaining channel-specific estimates.

Discrepancies between the Zebra BioDome version and the standard version were investigated by examining PCR and Luminex results, as well as histopathology and LBC results when available. Data on valid histopathology and/or LBC were available for 409 samples and both missing for 18 samples. A reference case was defined as HPV16/18/45/31/33/35/52/58 PCR positive for high-grade cervical intraepithelial neoplasia grade 2 or worse (CIN2+) on histopathology or LBC, and in addition HPV16 PCR positive for low-grade/equivocal lesions on histopathology or LBC. A reference control was defined as PCR high-risk HPV negative and neither histology nor LBC positive. Following these definitions, we identified 29 definite cases and 206 definite controls. 174 fell between the definitions of definite cases/controls.

Sensitivity analyses were carried out by restricting to HPV results of the study population of women within the age range of 25 to 49 years.

All analyses were carried out using Microsoft Excel, the Statistical Package for the Social Sciences (SPSS) (version 1.0.0.1089), and R Statistical Software (version 4.3.0).

## Results

### Repeatability

There was excellent agreement between the two replicates of ScreenFire HPV testing with the Zebra BioDome version on Powergene instrument 1 vs instrument 2 across all channels in the non-hierarchical analysis (Table 1). The replicate on instrument 2 showed a slight, but non-significant, increase in positivity compared to that on instrument 1 for the HPV31/33/35/52/58 channel. Repeatability was excellent also in the hierarchical analysis (not shown).

**Table 1:** Channel-specific repeatability of Zebra BioDome version of ScreenFire HPV RS assay, run on Powergene1 and Powergene2, amongst 426^#^ samples with valid results.

| Zebra BioDome ScreenFire HPV results: Powergene instrument 1/instrument 2 | +/+ (N) | -/+ (N) | +/- (N) | -/- (N) | Overall positive agreement [95% CI] | Overall negative agreement [95% CI] | McNemar p-value |
| --- | --- | --- | --- | --- | --- | --- | --- |
| HPV16 | 27 | 0 | 0 | 399 | 100% | 100% | NA<sup>*</sup> |
| HPV18/45 | 26 | 0 | 0 | 400 | 100% | 100% | NA<sup>*</sup> |
| HPV31/33/35/52/58 | 103 | 3<sup>a</sup> | 7<sup>b</sup> | 313 | 91.2%<br>[85.9%-96.4%] | 96.9%<br>[95.0%-98.8%] | 0.34 |
| HPV39/51/56/59/68 | 35 | 4<sup>c</sup> | 3<sup>d</sup> | 384 | 83.3%<br>[72.1%-94.6%] | 98.2%<br>[96.9%-99.5%] | 1.00 |
| **Agreement on overall HPV positivity (high-risk 13), irrespective of channel** | 175 | 5 | 6 | 240 | 94.1%<br>[90.7%-97.5%] | 95.6%<br>[93.1%-98.2%] | 1.00 |

<sup>#</sup>Excluding invalids on either replicate
<sup>\*</sup>McNemar test statistic cannot be derived because the disagreement is zero
<sup>a</sup>Three positives on PCR: one for HPV58 (signal strength=2), one for HPV35 (signal strength=2), one for HPV52 (signal strength=2)
<sup>b</sup>Three positives on PCR: one for HPV35 (signal strength=3), one for HPV31 (signal strength=1), one for HPV58 (signal strength=2)
<sup>c</sup>Two positives for HPV68 (signal strength=4 and 5) on PCR
<sup>d</sup>One positive for HPV68 (signal strength=3) and one positive for HPV51 (sign strength=1) on PCR

### Channel-specific agreement

The number of invalid results was similar when using Zebra BioDome and the standard assay version of the ScreenFire assay: 7 invalids by Zebra BioDome, 7 invalids by standard, 9 invalids by both. Among the 427 valid results by both, there was very good agreement between the Zebra BioDome version and the standard version across all channels in the non-hierarchical analyses (Table 2). Zebra BioDome showed a slight, but non-significant, increase in positivity compared to the standard version for the HPV16 and HPV31/33/35/52/58 channels.

**Table 2:** Channel-specific agreement (non-hierarchical) between Zebra BioDome (run on Powergene1) and the standard version of ScreenFire HPV RS assay amongst 427 samples with valid results by both versions.

| ScreenFire HPV results:<br>Zebra BioDome<br>(Powergene instrument<br>1)/Standard | +/+<br>(N) | -/+<br>(N) | +/-<br>(N) | -/-<br>(N) | Positive<br>agreement<br>with standard<br>as reference<br>[95% CI] | Negative<br>agreement with<br>standard as<br>reference [95%<br>CI] | McNemar<br>p-value |
| --- | --- | --- | --- | --- | --- | --- | --- |
| HPV16 | 25 | 0 | 2<sup>a</sup> | 400 | 100% | 99.5%<br>[98.8%-100%] | 0.48 |
| HPV18/45 | 25 | 1<sup>b</sup> | 1<sup>b</sup> | 400 | 96.2%<br>[88.8%-100%] | 99.8%<br>[99.3%-100%] | 1.00 |
| HPV31/33/35/52/58 | 104 | 1<sup>c</sup> | 7<sup>d</sup> | 315 | 99.0%<br>[97.2%-100%] | 97.8%<br>[96.2%-99.4%] | 0.08 |
| HPV39/51/56/59/68 | 38 | 5<sup>e</sup> | 0 | 384 | 88.4%<br>[78.8%-98.0%] | 100% | 0.07 |
| **Agreement on overall**<br>**HPV positivity (high-risk**<br>**13), irrespective of**<br>**channel** | 174 | 2 | 8 | 243 | 98.9%<br>[97.3%-100%] | 96.8% *<br>[94.6%-99.0%] | 0.11 |

<sup>a</sup>One of the two was positive for HPV16 on Luminex; the other was positive for HPV16 on PCR (signal strength=2)
<sup>b</sup>Additional positives on Zebra BioDome or standard version were positive for HPV18 on PCR (signal strength=2-3)
<sup>c</sup>One Positive for HPV31 on PCR (signal strength=2)
<sup>d</sup>Four positives on PCR: one for HPV52 (signal strength=1), one for HPV35 (signal strength=2), two for HPV58 (signal strength=2); three additional were positive for HPV53
<sup>e</sup>Three positives on PCR: two for HPV68 (signal strength= 4-5), one for HPV59 (signal strength=3)
\* Overall negative agreement was 96.8% [94.6%-99.0%] without sampling weights and was estimated to be 99.3% [98.8%-99.8%] after applying sampling weights for negatives.

Conversely, the standard version exhibited a slight, but non-significant, increase in positivity for the HPV39/51/56/59/68 channel. Additional positives detected by either version of the assay did not show specific cross-reactivity patterns with other high-risk or low-risk HPV types identified by PCR except for the possible chance finding that three of the seven additional positives by Zebra BioDome for HPV31/33/35/52/58 were positive for HPV53. The two additional positives by Zebra BioDome in the HPV16 channel were positive for HPV16 by either PCR or Luminex. Similar agreement in the channel-specific analysis was found when repeating the analysis only on the age range of 25 to 49 years and when using the results of the Zebra BioDome replicate on Powergene instrument 2 (not shown).

### Hierarchical agreement

There was excellent agreement between the Zebra BioDome version and the standard version of the ScreenFire assay also in the hierarchical analyses (Table 3), which is relevant for risk-based clinical management. Following the hierarchical distribution of HPV channels by the standard version, Zebra BioDome detected 25 out of 25 (100%) HPV16 positives; 24 out of 25 (96.0%) HPV18/45 positives in the absence of HPV16; 88 out of 90 (97.8%) HPV31/33/35/52/58 positives in the absence of HPV16, or 18/45; 35 out of 36 (97.2%) otherwise HPV39/51/56/59/68 positives. The overall agreement between the Zebra BioDome version vs the standard version was 97.2% (95% CI: 95.6%-98.8%) without sampling weights and 99.1% (95% CI: 98.6%-99.6%) with sampling weights (Table 3). Similar agreement in the hierarchical analysis was found when repeating the analysis using the results of the Zebra BioDome replicate performed on Powergene instrument 2 (not shown).

**Table 3:** Hierarchical agreement between Zebra BioDome and the standard version of ScreenFire HPV RS assay according to HPV risk groups amongst 427 samples with valid results by both versions.

| ScreenFire HPV results:<br>Zebra BioDome (Powergene<br>instrument 1)/Standard |  | Standard |  |  |  |  | Total for<br>Zebra<br>BioDome |
| --- | --- | --- | --- | --- | --- | --- | --- |
|  |  | HPV<br>16 | else<br>HPV<br>18/45 | else HPV<br>31/33/35/52/58 | else HPV<br>39/51/56/59/68 | else high-risk 13<br>negative |  |
| Zebra BioDome | HPV16 | 25 | 0 | 1 | 0 | 1 | 27 |
|  | column% | 100% | 0% | 1.1% | 0% | 0.4% | 6.3% |
|  | else HPV18/45 | 0 | 24 | 0 | 0 | 1 | 25 |
|  | column% | 0% | 96.0% | 0% | 0% | 0.4% | 5.9% |
|  | else<br>HPV31/33/35/52/58 | 0 | 1 | 88 | 0 | 6 | 95 |
|  | column% | 0% | 4.0% | 97.8% | 0% | 2.4% | 22.2% |
|  | else<br>HPV39/51/56/59/68 | 0 | 0 | 0 | 35 | 0 | 35 |
|  | column% | 0% | 0% | 0% | 97.2% | 0% | 8.2% |
|  | else high-risk 13<br>negative | 0 | 0 | 1 | 1 | 243 | 245 |
|  | column% | 0% | 0% | 1.1% | 2.8% | 96.8% * | 57.4% |
| Total for standard |  | 25 | 25 | 90 | 36 | 251 | 427 ** |

Highlighted in gray are concordant HPV risk group results.
\* Overall negative agreement was 96.8% without sampling weights and 99.3% after applying sampling weights for negatives.
\*\* Overall agreement was 97.2% without sampling weights and 99.1% after applying sampling weights for negatives.

Among the 29 reference cases, both the Zebra BioDome and the standard versions of the ScreenFire assay detected 27 cases. Both versions missed the same two (6.9%) cases: one positive for HPV16 and one positive for HPV31/33/35/52/58 by PCR.

Among the 206 reference controls, 13 (6.3%) were positive on both the Zebra BioDome and the standard versions of the ScreenFire assay. The Zebra BioDome version identified an additional three positives in the HPV31/33/35/52/58 channel.

## Discussion

Our results showed that the Zebra BioDome version of the ScreenFire HPV RS assay had excellent repeatability and agreement with the standard version of the assay, for each of the four type groups defined by the assay individually and hierarchically. Small differences in channel positivity and invalid results were seen but none were statistically significant.

The validation presented here aimed at comparing the novel Zebra BioDome version of ScreenFire HPV RS assay with the established standard version of the assay and assessing its repeatability on stored clinician-collected specimens under ideal laboratory conditions. A real-life trial of the clinical performance of Zebra BioDome post-shipping under realistic simpler laboratory conditions is beyond the scope of this work and is being addressed in ongoing field evaluation.

The Zebra BioDome version of the ScreenFire HPV RS assay was designed to manage the reality of HPV testing in LMICs, which is the need for accurate, rapid tests that allow single-day screening strategies for cervical cancer. As compared to the standard version of the ScreenFire HPV assay (8,9), Zebra BioDome seems to offer several advantages given its formulation with pre-aliquoted reagents and an anti-contamination component.

First, it eliminates the need for master mix preparation and aliquoting, hence reducing the number of manual pipetting steps. This translates into reduced hands-on time for assay preparation and reduced variability due to potentially unequal manually pipetted volumes (i.e., unequal volumes of primer mix and reaction mix combined during preparation of master mix, and/or unequal volumes of master mix dispensed in the reaction wells). The only manual pipetting step is the addition of 10 μL of lysate to the Zebra BioDome tube.

Second, the Zebra BioDome version reduces the risk of post-amplification contamination by sealing the products with the BioDome matrix material. Contamination of amplified PCR products in a molecular biology laboratory space is a critical concern, common to all PCR-based techniques (13). The assay’s high sensitivity means that even small amounts of DNA can be amplified, leading to a high number of copies post-amplification with a risk of leakage.

Contamination is very damaging because it compromises the trustworthiness of results due to false positives leading to incorrect interpretations and because it potentially invalidates the entire laboratory due to the need to decontaminate and repeat experiments, wasting time and resources. Separately from the work presented here, a few Zebra BioDome tubes were opened post-reaction, swabbed from the inside and re-run with ScreenFire HPV assay. They were found to have minimal to no amplification, thus demonstrating effective reduction of contamination with amplified products. By reducing the risk of contamination, the BioDome matrix increases the reliability of HPV results for clinical management. Besides the anti-contamination role, this matrix also acts as stabilizer of the Zebra BioDome formulation because it separates the layers of pre-aliquoted reagents (i.e., reaction mix and primer mix) avoiding their mixing and it creates a barrier from the air in the tube.

The formulation of Zebra BioDome may present a versatile and cost-effective solution for a wide range of DNA amplification techniques besides HPV, particularly at the table-top level. In fact, the pre-aliquoted reagents and anti-contamination components are applicable to other endpoint DNA amplification assays, potentially providing a broader application in research and diagnostic settings. This innovation could prove instrumental in addressing generic problems of molecular biology testing, such as assay preparation time and risk of post-amplification contamination, and in advancing the accessibility of molecular testing in all settings where cost and contamination control are critical.

The additional advantages of Zebra BioDome do not compromise those that are characteristic of the ScreenFire HPV RS assay in its original formulation (8,9). Most importantly, the risk-based hierarchy of HPV type channels because of the great difference in risk of cervical cancer associated with HPV types (9). Hence, in case of positivity for multiple HPV channel, clinical management is better directed following the hierarchical reading of the type groups: HPV16 positive, otherwise positive for HPV18 or HPV45 (if HPV16 was not detected), otherwise positive for HPV31/33/35/52/58 (if HPV16/18/45 were not detected), otherwise positive for HPV39/51/56/59/68, and otherwise negative. Like the standard version, the Zebra BioDome version allows for detection of HPV and a human DNA control directly from clinical specimens without the need for DNA extraction and is compatible with dry self-collected samples without requiring collection media. It also allows for flexibility to run up to 96 samples/controls without product wastage (i.e., the pre-aliquoted strips of 8-tubes can be used in multiples of 8 or cut as needed). These optimizations do not translate in additional costs compared to the standard version (approx. $6 per test for scale-up). For transport and storage, the space requirement is approximately unchanged while the temperature should be kept cold (recommended 2-8°C). A note of caution goes for the transportation conditions because high temperatures may cause disruption of the layers and invalidate the use of the product due to premature mixing of reagents, hence the importance of maintaining a cold chain and implementing temperature checks for this penetrable version of Zebra BioDome. In the future, a re-formulation may be considered to allow greater stability at warm temperatures and to remove the need of cold chain.

Following this positive evaluation, Zebra BioDome has been introduced at PAVE study sites using ScreenFire HPV RS assay (12). This field-evaluation will address the assay’s clinical accuracy and will evaluate its real-world use across a wide range of laboratory settings.

### Conclusions

In conclusion, the Zebra BioDome version of the ScreenFire HPV assay evaluated here has demonstrated accurate risk-based genotype grouping while simplifying the HPV testing process and reducing the risk of laboratory contamination. If validated in real-world settings, the assay will be an example of a rapid HPV extended genotyping solution for resource-limited settings as part of the ultimate public health goal to accelerate cervical cancer prevention. The formulation may also have potential application for other DNA amplification assays.

## Additional details

### Funding source

NCI Cancer Moonshot; NIH Intramural Research Program.

### Data availability statement

The data that support the findings of our study are available from the corresponding author upon reasonable request.

### Ethics statement

Both Nigerian and National Cancer Institute (NCI, US) institutional review boards (IRBs) (NCT 00804466) approved the original study protocol that included consent for specimen storage for future research.

### Authors contributions

Conceptualization, Mark Schiffman, Silvia de Sanjosé, Kanan T. Desai, Federica Inturrisi; methodology, Mark Schiffman, Kanan T. Desai, Silvia de Sanjosé, Federica Inturrisi, Didem Egemen; formal analysis, Kanan T. Desai, Federica Inturrisi, Didem Egemen; Itoju field investigation, Mark Schiffman, Kayode O. Ajenifuja, Clement A. Adepiti, Julia C. Gage, Nicolas Wentzensen; resources, Mark Schiffman, Kayode O. Ajenifuja, Clement A. Adepiti, Nicolas Wentzensen; data curation, Kayode O. Ajenifuja, Julia C. Gage, Clement A. Adepiti, Kanan T. Desai; laboratory evaluation, Amanda C. Hoffman, Casey L. Dagnall; writing—original draft preparation, Kanan T. Desai, Mark Schiffman, Silvia de Sanjosé, Federica Inturrisi; writing—review and editing, all. All authors have read and agreed to the published version of the article.

